# Treatment of COVID-19 Patients with Convalescent Plasma in Houston, Texas

**DOI:** 10.1101/2020.05.08.20095471

**Authors:** Eric Salazar, Katherine K. Perez, Madiha Ashraf, Jian Chen, Brian Castillo, Paul A. Christensen, Taryn Eubank, David W. Bernard, Todd N. Eagar, S. Wesley Long, Sishir Subedi, Randall J. Olsen, Christopher Leveque, Mary R. Schwartz, Monisha Dey, Cheryl Chavez-East, John Rogers, Ahmed Shehabeldin, David Joseph, Guy Williams, Karen Thomas, Faisal Masud, Christina Talley, Katharine G. Dlouhy, Bevin Valdez Lopez, Curt Hampton, Jason Lavinder, Jimmy D. Gollihar, Andre C. Maranhao, Gregory C. Ippolito, Matthew Ojeda Saavedra, Concepcion C. Cantu, Prasanti Yerramilli, Layne Pruitt, James M. Musser

## Abstract

**Background:** COVID-19 disease, caused by severe acute respiratory syndrome coronavirus 2 (SARS-CoV-2), has spread globally, and no proven treatments are available. Convalescent plasma therapy has been used with varying degrees of success to treat severe microbial infections for more than 100 years.

**Methods:** Patients (*n*=25) with severe and/or life-threatening COVID-19 disease were enrolled at the Houston Methodist hospitals from March 28 – April 14, 2020. Patients were transfused with convalescent plasma obtained from donors with confirmed SARS-CoV-2 infection and had been symptom free for 14 days. The primary study outcome was safety, and the secondary outcome was clinical status at day 14 post-transfusion. Clinical improvement was assessed based on a modified World Health Organization 6-point ordinal scale and laboratory parameters. Viral genome sequencing was performed on donor and recipient strains.

**Results:** At baseline, all patients were receiving supportive care, including anti-inflammatory and anti-viral treatments, and all patients were on oxygen support. At day 7 post-transfusion with convalescent plasma, nine patients had at least a 1-point improvement in clinical scale, and seven of those were discharged. By day 14 post-transfusion, 19 (76%) patients had at least a 1-point improvement in clinical status and 11 were discharged. No adverse events as a result of plasma transfusion were observed. The whole genome sequencing data did not identify a strain genotype-disease severity correlation.

**Conclusions:** The data indicate that administration of convalescent plasma is a safe treatment option for those with severe COVID-19 disease. Randomized, controlled trials are needed to determine its efficacy.

## Introduction

The coronavirus disease 2019 (COVID-19) pandemic has spread globally and caused massive loss of life and economic hardship. As of May 2, 2020, there were 3,494,671 confirmed cases and 246,475 deaths worldwide, and in the United States, there were 1,154,340 confirmed cases, and 67,447 deaths.^1^ The disease is caused by SARS-CoV-2, a highly transmissible coronavirus first identified in Wuhan, China.^2-4^ SARS-CoV-2 continues to spread in many countries,^5-9^ and despite aggressive research, no proven therapies have been described.

Treatment strategies for critically ill COVID-19 patients are lacking with only limited evidence available for a battery of anti-viral, antibiotic, and anti-inflammatory agents and aggressive supportive therapy. Multiple clinical trials are ongoing, including the repurposing of remdesivir, an anti-viral investigated to treat Ebola, and hydroxychloroquine (HCQ), an anti-malarial chloroquine derivative used to treat lupus and rheumatoid arthritis. There are early anti-COVID-19 efficacy data with remdesivir,^10^ and preliminary data supporting the use of HCQ, alone or in combination with azithromycin (AZM),^11^ has since been shown by larger controlled trials as misleading and potentially dangerous.^12^ New therapies are needed to improve outcomes for critically ill COVID-19 patients.

In convalescent plasma therapy, blood plasma from a recovered patient is collected and transfused to a symptomatic patient. The transfer of convalescent plasma is an old concept, having been used since at least 1918 when it was employed to fight the Spanish Flu pandemic.^13^ More recently, convalescent plasma was used with some reported success during the 2003 SARS pandemic,^14,15^ the 2009 influenza H1N1 pandemic,^16^ and the 2015 Ebola outbreak in Africa.^17^ Several small observational studies published during the COVID-19 pandemic suggest convalescent plasma is part of an effective treatment strategy for patients with severe disease.^18-21^ The first report describing administration of convalescent plasma to five patients early in the COVID-19 outbreak in Wuhan was recently published.^19^ Five critically ill patients received two, same-day infusions from five recovered healthy donors. In four of the five patients, inflammatory biomarkers decreased, A/a gradient improved, and all patients had improvement in pulmonary lesions based on computed tomography (CT) scan.^19^ A second study by Duan *et al*. reported improved clinical outcomes in 10 patients who received a single transfusion of convalescent plasma, and no adverse events reported.^18^ Two additional small case studies of five and six patients have since been published with similar findings.^20,21^ A more recent study by Zeng et. al. suggested that administration of convalescent plasma late in the disease course was ineffective for mortality reduction.^22^

We performed the present study to provide additional data on these initial clinical observations of patients’ clinical course and subsequent improvement after receiving convalescent plasma therapy for COVID-19. We transfused 25 COVID-19 patients with severe and/or life-threatening disease at the Houston Methodist hospitals, a large, quaternary-care hospital system that serves metropolitan Houston, Texas (~7 million people).^23^ Patients were transfused once with 300 mL of convalescent plasma. The therapy was well-tolerated and no transfusion-related adverse events were observed. At day 7 post-transfusion, nine of 25 patients (36%) had improvement in the assessed clinical endpoints. By 14 days post-transfusion, 19 patients (76%) had improved or been discharged. Although our study has limitations, the data indicate that transfusion of convalescent plasma is a safe treatment option for those with severe COVID-19 disease.

## Methods

This study was conducted at the Houston Methodist hospitals from March 28, 2020, through April 28, 2020, with the approval of the Houston Methodist Research Institute ethics review board and with informed patient or legally-authorized representative consent. Patients were treated either under emergency investigational new drug (eIND) or investigational new drug (IND) applications approved by the U.S. Food and Drug Administration (https://www.fda.gov/medical-devices/emergency-situations-medical-devices/emergency-use-authorizations). Approval to treat the first patient by eIND was granted on March 28, 2020. The IND application was approved on April 3, 2020.

### Patients

COVID-19 patients in the Houston Methodist hospitals were considered for enrollment in this trial. SARS-CoV-2 infection was confirmed by RT-PCR. Patients were eligible if they had severe and/or life-threatening COVID-19 disease.^24^ Severe disease was defined as one or more of the following: shortness of breath (dyspnea), respiratory rate ≥ 30/min, blood oxygen saturation ≤ 93%, partial pressure of arterial oxygen to fraction of inspired oxygen ratio < 300, and/or pulmonary infiltrates > 50% within 24 to 48 hours. Life-threatening disease was defined as one or more of the following: respiratory failure, septic shock, and/or multiple organ dysfunction or failure. Clinical data for patients was obtained from the hospital electronic medical record.

### Definition of Clinical Disease Severity

Clinical severity for the purposes of outcome assessment was scored based on a modified 6-point clinical scale used by the WHO R&D Blueprint group and others.^25,26^ Patients were assigned a clinical status at baseline (day zero, date of transfusion) and evaluated at days 0, 7, and 14. The 6-point scale is as follows: 1, discharged (alive); 2, hospitalized, not requiring supplemental oxygen but requiring ongoing medical care (for COVID19 or otherwise); 3, hospitalized, requiring low-flow supplemental oxygen; 4, hospitalized, on non-invasive ventilation or high-flow oxygen devices; 5, hospitalized and on invasive mechanical ventilation or extracorporeal membrane oxygenation (ECMO); and 6, death.

### Convalescent Plasma Donors

Convalescent plasma was obtained by apheresis using the Trima Accel automated blood collection system (Terumo BCT). Plasma (600 mL) was collected from each donor and divided into two 300 mL units. Each donor had a documented history of laboratory-confirmed SARS-CoV-2 infection based on a positive RT-PCR test result. All plasma was donated by recovered and healthy COVID-19 patients who had been asymptomatic for 14 or more days. Donors were between 23-67 years old. All donors provided written informed consent and tested negative for SARS-CoV-2 by RT-PCR. If eligible according to standard blood donor criteria, donors were enrolled in a frequent plasmapheresis program. Donors were negative for anti-HLA antibodies, hepatitis B, C, HIV, HTLV I/II, Chagas disease, WNV, Zika virus, and syphilis per standard blood banking practices.

### RT-PCR Testing for SARS-CoV-2 Infection

Symptomatic patients with a high degree of suspicion for COVID-19 disease were tested in the Molecular Diagnostics Laboratory at Houston Methodist Hospital using a validated assay applied for under Emergency Use Authorization (EUA) from the U.S. Food and Drug Administration (https://www.fda.gov/medical-devices/emergency-situations-medical-devices/emergency-use-authorizations). The assay follows the protocol published by the World Health Organization^27^ and uses a 7500 Fast Dx instrument (Applied Biosystems) and 7500 SDS software (Applied Biosystems). Testing was performed on nasopharyngeal or oropharyngeal swabs immersed in universal transport media (UTM), bronchoalveolar lavage fluid, or sputum treated with dithiothreitol (DTT).

### SARS-CoV-2 ELISA

Costar 96-well assay plates (Corning) were coated with either SARS-CoV-2 spike (S protein) ectodomain (ECD) or SARS-CoV-2 spike receptor binding domain (RBD) (50 μL at 2 μg/mL in PBS) overnight at 4°C. **Detailed methods on protein purification and ELISAs can be found in supplemental methods**. Plates were blocked with 2% milk in PBS at room temperature (RT) for 2 hrs and washed 3X with PBST (PBS with 0.1% Tween20). Plasma or mAb was serially diluted in 50 μL/well across the entire 96-well plate. Negative plasma control was included on each antigen plate. mAb CR3022 was used as a positive control. CR3022 is a neutralizing antibody originally cloned from a convalescent SARS patient that targets the RBD of SARS-CoV^28^ and binds to the RBD of SARS-CoV-2 with a binding affinity of 6.3 nM.^29^ Binding was performed at RT for 1 hr. Plates were washed and anti-human IgG Fab HRP (Sigma A0293, 1:5000) was added to the plate (50 μL), and incubated at RT for 30 min. Plates were washed 3X with PBST, ELISA substrate (1-step Ultra TMB, Thermo Scientific cat# 34028) was added, plates were developed for 1 min for RBD and 5 min for spike ECD, and the reaction was stopped with 50 μL of H_2_SO_4_. Plates were read at 450 nm absorbance. Three-fold serial dilutions from 50 to 4050 were analyzed. Titer was defined as the last dilution showing an optical density greater than a multi-plate negative control average plus six standard deviations.

### SARS-CoV-2 Genome Sequencing and Analysis

Libraries for whole viral genome sequencing were prepared according to version 1 ARTIC nCoV-2019 sequencing protocol (https://artic.network/ncov-2019). Long reads were generated with the LSK-109 sequencing kit, 24 native barcodes (NBD104 and NBD114 kits), and a GridION instrument (Oxford Nanopore). Short reads were generated with the NexteraXT kit and a MiSeq or NextSeq 550 instrument (Illumina). Whole genome alignments of consensus viral genome sequence generated from the ARTIC nCoV-2019 bioinformatics pipeline were trimmed to the start of orf1ab and the end of orf10 and used to generate a phylogenetic tree using RAxML (https://cme.h-its.org/exelixis/web/software/raxml/index.html) to determine viral clade. Trees were visualized and annotated with CLC Genomics Workbench v20 (Qiagen).

## Results

### Overview of Patient Characteristics

Twenty-five patients with severe and/or life-threatening COVID-19 disease were enrolled in the study from March 28 – April 14, 2020. Patients ranged in age from 19 to 77 years (median 51, interquartile range [IQR] 42.5 to 60), and 14 were female (**Table 1**). The median BMI was 30.4 kg/m^2^ (IQR 26.5 to 37) and the majority (22/25, 88%) had no smoking history. Many patients (16/25, 64%) had one or more underlying chronic conditions, including diabetes (10 patients), hypertension (9 patients), hyperlipidemia (5 patients), and gastrointestinal reflux disease (GERD, 4 patients). The majority of patients (19 of 25, 76%) enrolled in the study had O-positive blood type. Bacterial or viral co-infections were identified in five patients (**Table 1**).

**Table 1:** Demographics and Clinical Characteristics of Patients with COVID-19 Disease who Received Convalescent Plasma.

| Patient | Sex | Age | Weight,<br>kg | BMI<br>(kg/m<sup>2</sup>) | Smoking<br>History | Blood<br>Type | Co-Infections | Coexisting Chronic<br>Diseases |
| --- | --- | --- | --- | --- | --- | --- | --- | --- |
| 1 | F | 39 | 90 | 34 | Never | O pos | None | DM2 |
| 2 | F | 63 | 104 | 38 | Never | O pos | None | DM2, HTN, HLP,<br>GERD |
| 3 | F | 48 | 63 | 23 | Never | O pos | None | None |
| 4 | M | 57 | 96 | 29 | Never | O pos | None | None |
| 5 | F | 38 | 99 | 35 | Never | O pos | Influenza B | DM2, HTN, GERD |
| 6 | M | 46 | 133 | 32 | Former | O pos | MSSA PNA | DM2 |
| 7 | M | 51 | 94 | 32 | Former | A pos | None | DM2 |
| 8 | M | 74 | 84 | 27 | Never | A pos | VAP: MSSA &<br>GAS | DM2, HTN, CKD |
| 9 | F | 55 | 73 | 26 | Never | O pos | None | None |
| 10 | F | 19 | 113 | 49 | Never | O pos | Enterococcus<br>BSI | None |
| 11 | F | 22 | 91 | 40 | Never | O pos | None | Asthma |
| 12 | F | 46 | 65.8 | 24.9 | Never | O pos | None | None |
| 13 | M | 61 | 88 | 30 | Unknown | O pos | None | None |
| 14 | F | 49 | 101 | 31.9 | Never | O pos | None | GERD, HTN |
| 15 | M | 29 | 126 | 44 | Never | O pos | None | None |
| 16 | F | 30 | 94.7 | 38.2 | Never | O pos | None | Post-partum,<br>hypothyroidism |
| 17 | F | 54 | 79 | 30 | Never | O pos | None | HTN |
| 18 | M | 56 | 102 | 40 | Never | O pos | None | HTN, HLP |
| 19 | M | 60 | 81.6 | 32 | Never | O pos | None | DM2, HLD |
| 20 | F | 77 | 95 | 36 | Never | O pos | None | HTN, DM2 |
| 21 | F | 60 | 65 | 23 | Never | O neg | None | None |
| 22 | F | 77 | 86.5 | 29.8 | Never | A pos | GAS | Atrial fibrillation, DM2,<br>HLD |
| 23 | M | 60 | 85 | 30.4 | Never | O pos | None | DM2, HLD, HTN |
| 24 | M | 54 | 72 | 25 | Never | B pos | None | HLD |
| 25 | M | 50 | 58 | 22.6 | Never | B pos | None | None |

Abbreviations: F, Female; M, Male; pos, positive; VAP, ventilator-associated pneumonia; MSSA, methicillin-susceptible *Staphylococcus aureus*; GAS, group A Streptococcus; PNA, pneumonia; BSI, bloodstream infection; DM2, diabetes mellitus 2; HTN, hypertension; GERD, gastrointestinal reflux disease; HLD, hyperlipidemia; AF, atrial fibrillation; CKD, chronic kidney disease; None, indicates no infection identified.

### Donor Characteristics

The characteristics of the donors of convalescent plasma are shown in **Table 2**. A total of nine donors provided plasma that was used to transfuse COVID-19 patients; two donors gave plasma on multiple occasions. The donors ranged in age from 23 to 67 years, and 56% (5/9) were males. On average, the donors gave plasma 26 days (range 19-33) from their symptom start date and 21 days (range 13 to 27 days) from their initial positive RT-PCR specimen collection date. Although all donors had been symptomatic, only one was ill enough to require hospitalization. To assess antibody titers, we used two ELISAs, one based on recombinant purified ectodomain (ECD) of the spike protein and the second using recombinant receptor binding domain (RBD) of the spike protein. The titers of the convalescent plasma used for transfusion ranged from 0 to 1350 for the RBD and ECD domains (**Figure 1 and Table S1**).

**Table 2.**
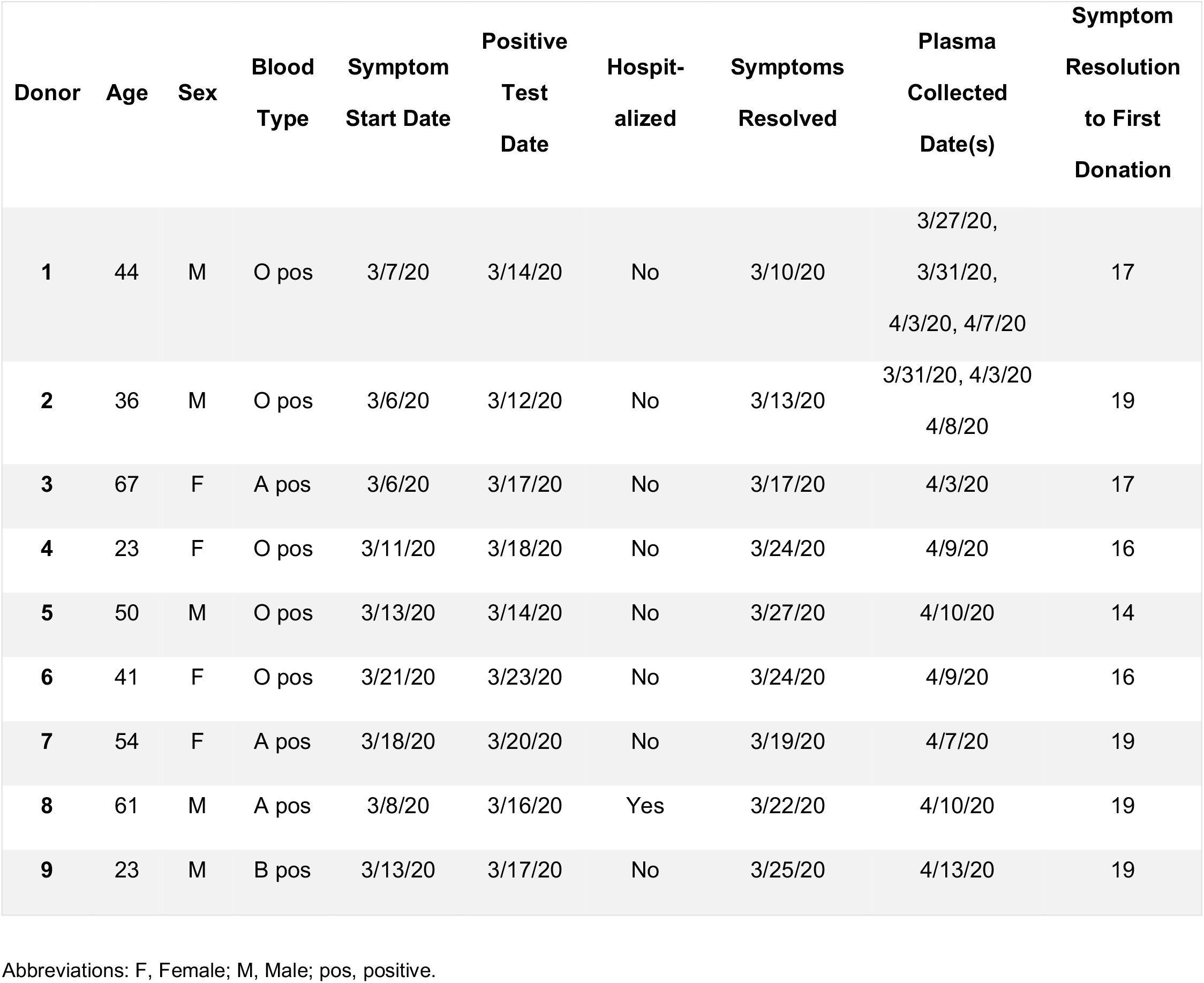
Characteristics of Convalescent Plasma Donors.

**Figure 1.**
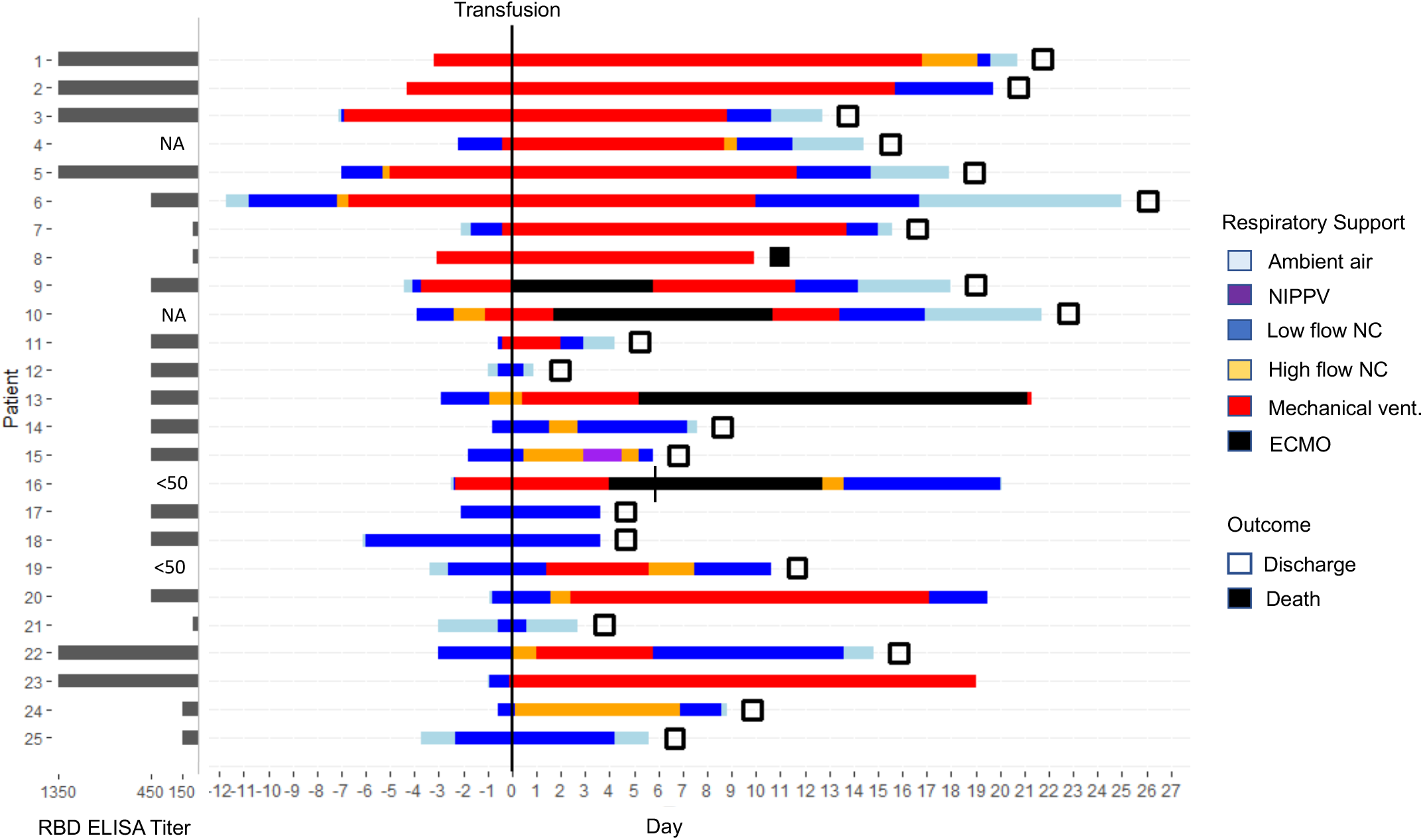
Respiratory support status, clinical score, patient outcomes (discharge/death), and RBD titer of transfused plasma in 25-patient cohort. Respiratory support requirements for the duration of hospitalization are color coded per the key. Discharge or death are indicated by open and filled squares, respectively. Patient 16 was given a second transfusion on day 6, indicated by a vertical line. The convalescent plasma titers for the RBD domain of the SARS-CoV-2 spike protein are indicated to the left. NIPPV, non-invasive, positive-pressure ventilation; NC, nasal cannula; ECMO, extracorporeal membrane oxygen.

### Transfusion of Severe COVID-19 Patients with Convalescent-Phase Donor Plasma

The median time from symptom onset to hospitalization was 6 days [IQR 4 to 8 days] (**Table 3**). The majority of patients received concomitant anti-inflammatory treatments within five days of the plasma transfusion, including tocilizumab and steroids. Most received other investigational treatments, including courses of HCQ and AZM, ribavirin, and/or lopinavir/ritonavir, and two patients received remdesivir (**Table 3**). All patients required oxygen support prior to transfusion (**Figure 1**), including 12 patients on mechanical ventilation, 10 on low-flow oxygen, and 3 on high-flow oxygen. One patient (patient 9) was placed on ECMO on the day of transfusion prior to transfusion. More than half (13/25, 52%) had acute respiratory distress syndrome (ARDS)^30^ at the time of transfusion (**Table 3**). The median time from symptom onset to transfusion was 10 days [IQR, 7.5 to 12.5], and from hospitalization to transfusion was 2 days [IQR, 2 to 4] (**Table 3**). All patients received one 300-mL dose of convalescent-phase plasma, and one patient received a second transfusion six days after the initial transfusion. Clinical outcomes and laboratory parameters were assessed at days 0, 7, and 14 post-transfusion.

**Table 3.** Disease Course and Additional Treatments of Patients Receiving Convalescent Plasma.

| **No.** | **Symptom**<br>**Onset to**<br>**Admission**<br>**(d)** | **Symptom**<br>**Onset to**<br>**Positive SARS**<br>**Test (d)** | **Admission to**<br>**Transfusion**<br>**(d)** | **Complications**<br>**Prior to**<br>**Transfusion** | **Anti-**<br>**Inflammatory**<br>**Treatments** | **Antiviral**<br>**Treatments** | **Length of**<br>**Hospital Stay**<br>**(d)** | **Post-Transfusion**<br>**Length of**<br>**Hospital Stay (d)** |
| --- | --- | --- | --- | --- | --- | --- | --- | --- |
| 1 | 7 | 3 | 1 | ARDS | Tocilizumab | HCQ, RBV | 24 | 21 |
| 2 | 7 | 9 | 4 | ARDS, CRRT | Interferon,<br>Steroids | HCQ, AZM,<br>RBV | 24 | 20 |
| 3 | 8 | 3 | 6 | ARDS | Tocilizumab,<br>Steroids | HCQ, RBV,<br>LPVr | 20 | 13 |
| 4 | 8 | 9 | 2 | ARDS | Tocilizumab,<br>Steroids | HCQ, AZM,<br>RBV | 17 | 15 |
| 5 | 3 | 4 | 7 | ARDS | None | HCQ, AZM,<br>RBV | 25 | 18 |
| 6 | 3 | 4 | 13 | ARDS | Tocilizumab | HCQ, AZM,<br>RBV | 37 | NA |
| 7 | 3 | 3 | 2 | ARDS | Tocilizumab | HCQ, LPVr | 20 | 16 |
| 8 | 4 | 5 | 3 | ARDS | Steroids | HCQ, RBV,<br>LPVr | 13 | 10 |
| 9 | 4 | 4 | 4 | ARDS, CRRT,<br>ECMO (VV) | Tocilizumab,<br>Steroids | HCQ, RBV | 22 | 18 |
| 10 | 6 | 10 | 5 | ARDS | Tocilizumab | HCQ, AZM,<br>RBV, LPVr,<br>remdesivir | 28 | 22 |
| 11 | 5 | 3 | 1 | ARDS | Steroids | HCQ, AZM,<br>RBV | 5 | 4 |
| 12 | 10 | 6 | 2 | None | None | HCQ, AZM | 2 | 1 |
| 13 | 5 | 6 | 3 | None | Tocilizumab | HCQ, AZM,<br>RBV | NA | NA |
| 14 | 12 | 6 | 1 | None | Tocilizumab,<br>Steroids | HCQ, AZM,<br>RBV | 9 | 8 |
| 15 | 7 | 8 | 2 | None | None | HCQ, AZM,<br>RBV | 8 | 6 |
| 16 | 8 | 3 | 2 | ARDS | Tocilizumab,<br>Steroids | HCQ, AZM,<br>RBV | NA | NA |
| 17 | 4 | 4 | 2 | None | None | HCQ, AZM | 6 | 4 |
| 18 | 8 | 8 | 6 | None | None | HCQ, AZM | 10 | 4 |
| 19 | 6 | 6 | 3 | None | Tocilizumab | HCQ, AZM | 14 | 11 |
| **20** | 3 | 4 | 1 | None | None | HCQ, RBV,<br>AZM,<br>remdesivir | NA | NA |
| **21** | 8 | 8 | 3 | None | None | HCQ, RBV | 6 | 3 |
| **22** | 4 | 4 | 2 | None | Steroids | HCQ, AZM,<br>RBV | 18 | 15 |
| **23** | 14 | 1 | 2 | ARDS | Tocilizumab,<br>Steroids | HCQ, AZM | NA | NA |
| **24** | 9 | 6 | 2 | None | Tocilizumab | HCQ, AZM | 10 | 9 |
| **25** | 11 | 11 | 3 | None | Tocilizumab | HCQ, AZM | 9 | 6 |

Abbreviations: ARDS, acute respiratory distress syndrome; CRRT, cardiac rapid response team; ECMO (VV), extracorporeal mechanical oxygenation (veno-venous); IFN, Interferon; HCQ, hydroxychloroquine; AZM, azithromycin; RBV, ribavirin; LPVr, lopinavir/ritonavir; NA, still hospitalized at day 14 post-transfusion (study endpoint).

### Outcomes

The primary clinical endpoint of the study was safety. No adverse events attributed to plasma transfusion occurred within 24 hours after transfusion. One patient developed a morbilliform rash one day post-transfusion that lasted for several days. Punch biopsy findings were compatible with an exanthematous drug eruption, and classic histologic findings of serum sickness (leukocytoclasic vasculitis) were not seen. Two patients developed deep-vein thrombosis (DVT) four and eight days after transfusion, and one patient developed a DVT and a pulmonary embolism four days post-transfusion. The observed thrombotic complications are consistent with findings reported for COVID-19 patients.^31^ The secondary endpoint was an improvement in the modified 6-point WHO ordinal scale at day 14 post-transfusion including discharge from the hospital (**Table S2**). At day 7 post-transfusion, nine patients (36%) improved from baseline, 13 (52%) had no change, and three deteriorated (**Figure 2**). Seven of the nine improved patients (28%) had been discharged. By day 14 post-transfusion, 19 (76%) patients improved from baseline: an additional four patients were discharged, eight patients improved from baseline, three patients remained unchanged, three had deteriorated, and one patient died from a condition not caused by plasma transfusion (**Figure 2 and Table S2**). The average overall length of hospital stay was 14.3 days (range 2 to 25 days). The average post-transfusion length of hospital stay was 11 days (range 1 to 21 days) (**Table 3**).

**Figure 2.**
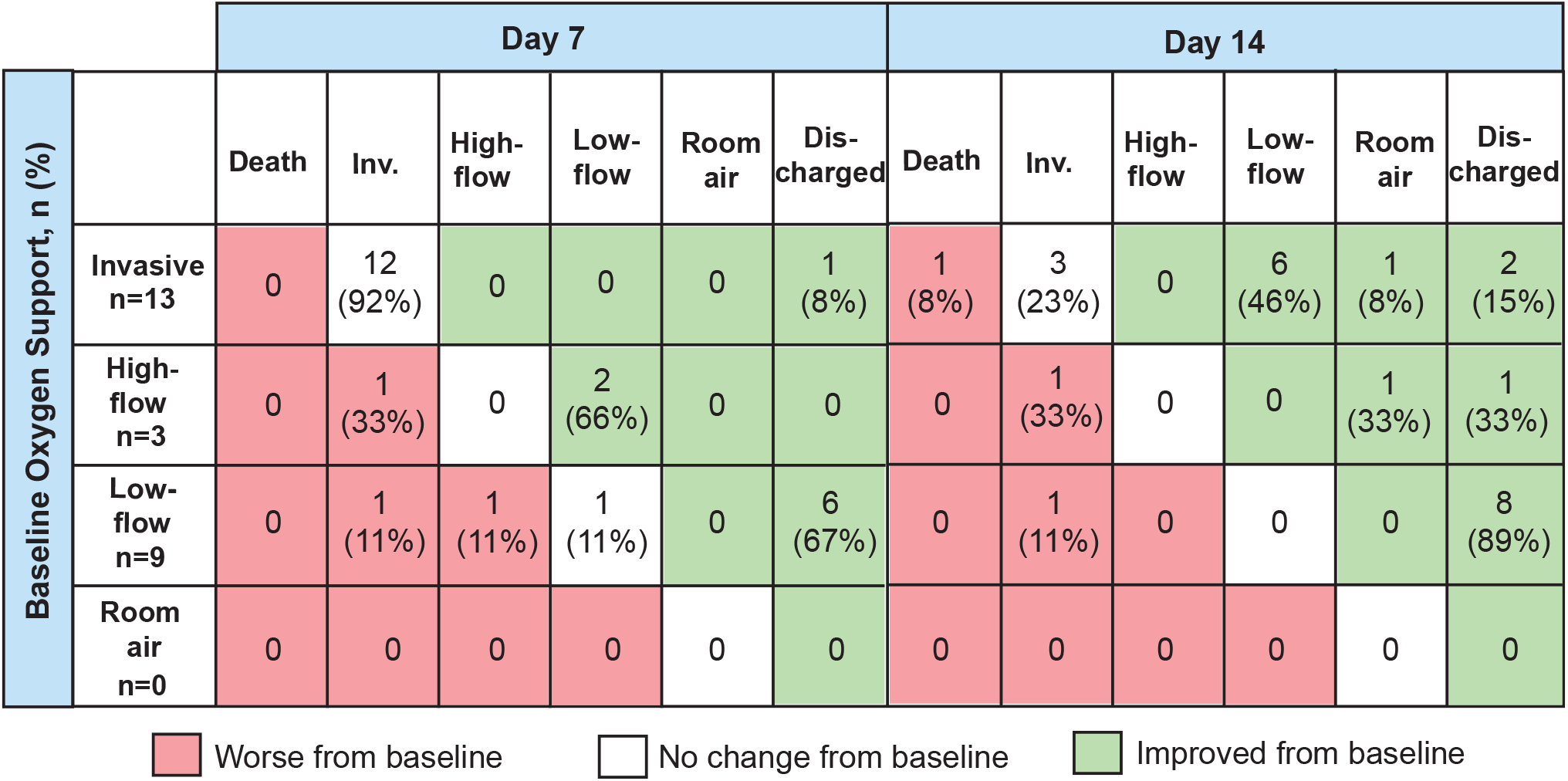
Clinical outcomes at day 7 and 14 post-transfusion. The table shows the distribution of patients on low-flow, high-flow, invasive, or no oxygen support at days 0 (day of transfusion), 7, and 14. By day 7 post-transfusion, 36% (9/25) of patients had improved from baseline; 76% (19/25) of patients improved by day 14 post-transfusion.

### Laboratory results

Laboratory results were assessed for parameters associated with inflammation and liver function. The median value for C-reactive protein decreased in our cohort from 14.66 mg/dL at day 0 to 2.9 mg/dL and 0.45 mg/dL at days 7 and 14 post-transfusion, respectively (**Table 4**). There was a trend toward increasing ferritin by day 3, which tended to decrease by day 7. No significant increases in liver enzymes were noted (**Table 4 and Table S3**).

**Table 4.**
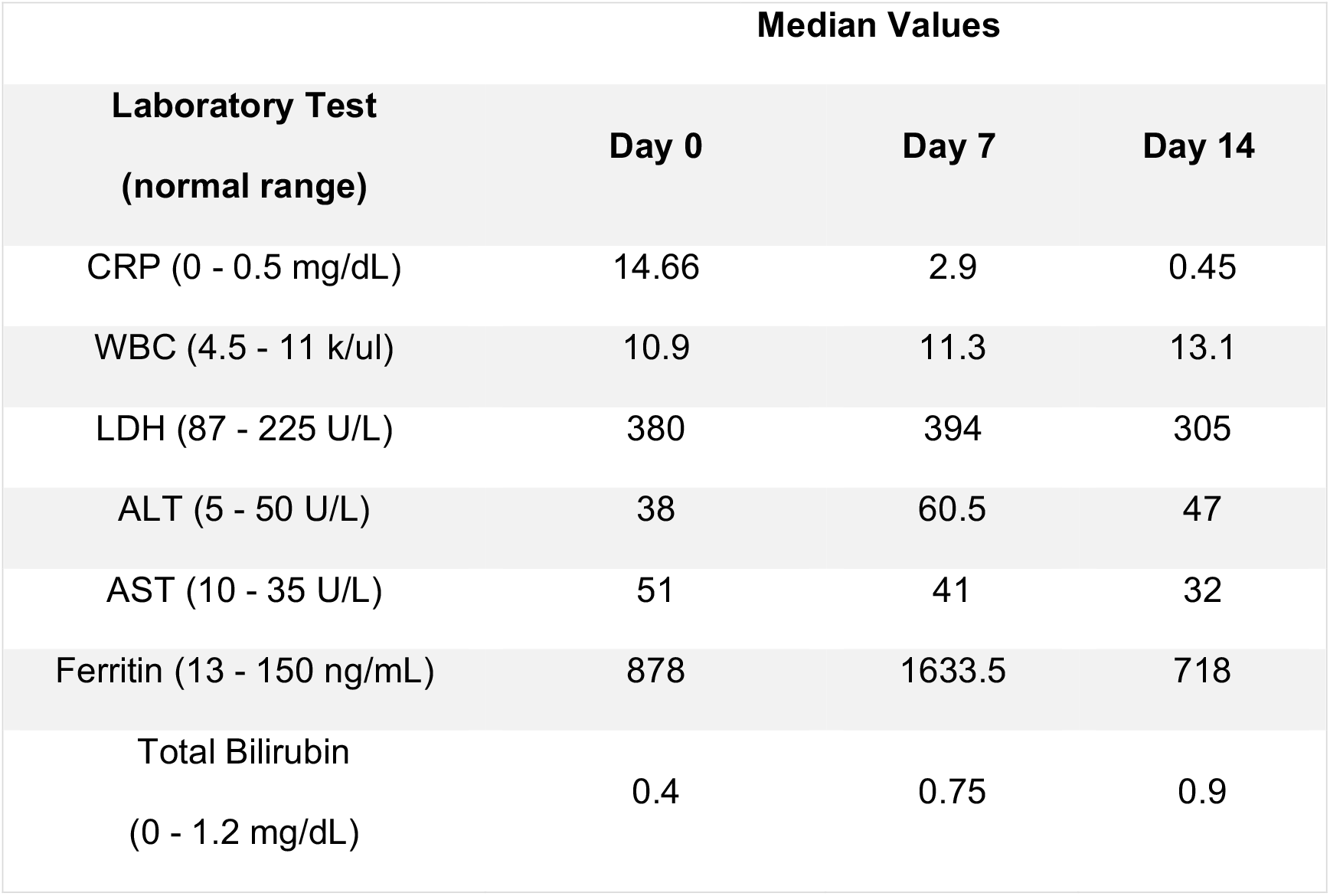
Median Laboratory Values of Plasma Recipients at Days 0, 7, and 14 Post-Transfusion.

### Viral Genome Sequencing of SARS-CoV-2 Strains from Recipients and Donors

A recent analysis of the genomic heterogeneity of the SARS-CoV-2 virus strains circulating in Houston, Texas, early in the pandemic showed that the predominant clades isolated were A2a, B, and B1.^32^ Amino acid polymorphisms, especially in the spike protein, can potentially alter the character of the antibody response and virulence profile of the virus.^33-36^ Therefore, we sequenced the genomes of the SARS-CoV-2 virus strains infecting donors and recipients to assess the magnitude of nucleotide and amino acid mismatch between the viral genotype of donors and plasma recipients. Of the 34 patients and donors, we were able to analyze all plasma recipient genotypes and four donor genotypes. Overall, there were few polymorphisms in the sequenced viruses, and there was no correlation between infecting strains and disease severity (**Figure S1**). Analysis of the first four donors found that, in general, donor and recipient S proteins matched when their SARS-CoV2 isolates were from the same clade (**Figure S1**). This is primarily a result of a D614G amino acid change in S protein that defines the clade A2a.^32,37^ However, there are at least three instances of an additional amino acid change in the S2 domain of the S protein,^34-36^ one in a donor (M731I) and two in recipients (S967R and L1203F) (**Figure S1**).

## Discussion

Our study was performed to evaluate the safety and potential benefit of transfusing convalescent plasma to patients with severe COVID-19 disease. To date, this is the largest cohort assessed for outcomes pertaining to convalescent plasma transfusion for COVID-19. Of our 25 patients, nine showed improvement by day 7, and by day 14 post-transfusion, 19 patients showed improvement, as assessed by discharge or at least a 1-point improvement on a modified clinical scale. Several case studies investigating the use of convalescent plasma to treat severe COVID-19 have recently been published^18-22^ and the overall findings presented herein are consistent with these reports.

Convalescent plasma therapy has been administered on the front lines during emergencies, and we and others recognize the need for controlled clinical trials to determine its therapeutic efficacy.^14,15,19,38,39^ The timing of the transfusion post-symptom onset, the number of transfusions, the volume and its adjustment based on BMI, donor antibody titers, and other parameters need to be evaluated to optimize this therapy. For example, some studies have observed that the sooner after the onset of symptoms that the transfusion was administered, the better the outcomes.^14,15,39^ Variability existed among our cohort with respect to symptom onset and severity of illness.

The anti-SARS-CoV2 anti-spike protein IgG titers varied significantly among individual donors, as assessed by ELISA (**Table S1**). Early in the study period, ELISA titers were not available, and thus, transfusions were given solely on the basis of ABO compatibility. Among the five patients who received plasma from a donor with an anti-RBD IgG titer of ≤50, one is deceased, and one was placed on ECMO. The patient placed on ECMO received a second dose of convalescent plasma confirmed to have high IgG titer prior to transfusion. The patient was eventually extubated and weaned off ECMO. Regardless, at this time, no clear correlation between ELISA IgG titer and patient outcomes can be established in this small patient cohort. In addition, more studies are needed to better understand why donors present with a range of anti-spike antibody titers, and whether there is a correlation between donor disease presentation and antibody titers. Additional studies are underway to better understand the correlation between anti-SARS-CoV-2 antibody titers and virus neutralization.

The results from our study support the existing data from the COVID-19 literature that point to underlying medical conditions, such as obesity, type 2 diabetes, and hypertension, playing a large role in patients’ COVID-19 disease course and outcomes.^40-42^ Sixty-eight percent (17/25) of transfused patients in this study had a BMI in the obese category and 84% were considered overweight.

A confounding variable in many convalescent plasma studies is the addition of other treatment regimens, such as antivirals and anti-inflammatory compounds. Adjunct therapies hinder the ability to draw definitive conclusions regarding the contribution of the convalescent plasma. In our study, all 25 patients received HCQ and AZM, as these were reported to have beneficial effects early in the pandemic.^11^ Subsequent larger and more controlled studies determined that this combination has no benefits to patients, and in fact, could be harmful.^12^ Many (68%) of our patients were also administered oral ribavirin. Despite inconclusive data on ribavirin’s efficacy in the treatment of SARS during the 2003 epidemic,^43^ proven safety and ready availability supported its use in the treatment of our COVID-19 patients. Two patients received remdesivir, which was recently shown to modestly reduce recovery time.^10,44^ Anti-inflammatory compounds, such as the IL-6 inhibitor tocilizumab and methylprednisolone, were administered per institutional protocols within five days of the plasma transfusion to 72% of our cohort. Tocilizumab was recently shown to reduce mortality in a retrospective analysis of 20 severe COVID-19 patients.^45^ Because convalescent plasma therapy is typically performed in emergency situations for the very ill, it is difficult to assess its benefits as a standalone treatment. A blinded, randomized controlled trial is currently being considered.

The patient outcomes in our study are similar to those recently published describing treating COVID-19 patients with remdesivir on a compassionate-use basis.^10^ In that review, patients were prescribed a 10-day course of remdesivir with follow up for 28 days or until discharge or death. Both study cohorts included patients who required invasive ventilation, including 35 of 53 (66%) of remdesivir patients compared to 17 of 25 (68%) of the patients in our study. Clinical improvement was less frequent among patients who received invasive ventilation at any time or were 70 years of age or older. In the remdesivir study, 36 of 53 patients (68%) showed clinical improvement at follow up (median time to follow up, 18 days), while 19 of 25 patients (76%) receiving convalescent plasma improved by day 14 post-transfusion. These data suggest that treatment with convalescent plasma and remdesivir resulted in similar outcomes among patients based on oxygenation requirements and age. The mortality difference between the cohorts cannot be compared as the remdesivir cohort represented an older population (median age of 64 years, versus 51 years in our study), where the risk of death was greater at baseline. Delays in obtaining remdesivir on a compassionate use basis (12 days from symptom onset) may have artificially extended the cohort’s opportunity to demonstrate clinical improvement and does not reflect the eligibility criteria for any ongoing clinical trials.^46-48^ Clinical outcomes data to inform timing of therapeutic interventions like remdesivir or convalescent plasma are lacking.

We analyzed the genomes of the infecting SARS-CoV-2 strain from both the donors and recipients. One could conceive of a situation in which the donor genotype of the SARS-CoV-2 infecting strain was matched with the genotype of the patient’s strain to maximize potential immune benefit. We found few differences in the inferred amino acid sequences of the plasma donor and recipient strains, and no association between disease severity and infecting strain genotype.

The majority of the donors and plasma recipients in our study had type O blood (25/34, 74%). Our initial donors, who donated repeatedly, were blood type O. Since ABO-compatibility was a requirement for recipient selection early in the study, many of our early recipients were also type O. Zhao *et al*. have reported that of the 2,173 patients analyzed in their study of COVID-19 patients in China, the majority had type A blood.^49^ More studies are needed to determine if this association holds true in geographically-distinct areas of infection. Regardless, our data do not reflect a higher rate of blood type A in COVID-19 patients.

### Limitations

As with the great majority of the studies using convalescent plasma to treat severe infections, our study has several important limitations. First, the study was a small case series and no control group was included. Thus, it is not clear if the 25 patients given convalescent plasma would have improved without this treatment. Second, all patients were treated with multiple other medications, including antiviral and anti-inflammatory agents. Thus, we cannot conclude that the patient outcomes were solely due to administration of convalescent plasma. Third, 24 of the 25 patients received only one transfusion of plasma. Whether treatment with multiple transfusions on one or more days would be a more effective regimen is not clear. An expanded donor pool providing higher-titer convalescent plasma would allow for dose escalation studies. Fourth, many patients had severe COVID-19 disease. It is possible that transfusion of convalescent plasma earlier in the course of disease or in patients with less severe symptoms would be a better approach. Fifth, our plasma donors had a range of anti-S protein IgG titers. Several patients were transfused with plasma with very low titer of anti-S protein antibody. Sixth, the small number of patients treated, coupled with the experimental design, did not permit us to determine if this therapy significantly reduces mortality or other measures of disease outcome. Finally, while this study assessed outcomes at days 7 and 14 post-transfusion, it is important to note that at the time of this writing, all but two of the surviving patients that were intubated had been extubated. Similarly, all patients that were on ECMO had been weaned, and 20 of the 25 patients had been discharged.

## Concluding Statement

Outcomes from this case series of 25 patients indicates that administration of convalescent plasma is a safe treatment option for those with severe COVID-19 disease.

## Data Availability

There is no data availability issue.

## Acknowledgments

We are deeply indebted to our many generous volunteer plasma donors for their time, their gift, and their solidarity. We thank Drs. Jessica Thomas and Zejuan Li, Erika Walker, the very talented and dedicated molecular technologists, and the many labor pool volunteers in the Molecular Diagnostics Laboratory for their dedication to patient care. We thank the many donor center and blood bank phlebotomists and technologists for their dedication to donor and blood safety. We thank Kathryn Stockbauer for editorial assistance. We also thank Brandi Robinson, Harrold Cano, and Cory Romero for technical assistance. We thank Susan Miller and Mary Clancy for consistent and thorough advice. We are indebted to Drs. Marc Boom and Dirk Sostman for their support, and to many very generous Houston philanthropists for their tremendous support of this ongoing project, including but not limited to anonymous, Ann and John Bookout III, Carolyn and John Bookout, Ting Tsung and Wei Fong Chao Foundation, Ann and Leslie Doggett, Freeport LNG, the Hearst Foundations, Jerold B. Katz Foundation, C. James and Carole Walter Looke, Diane and David Modesett, the Sherman Foundation, and Paula and Joseph C. “Rusty” Walter III. Dr. Jason S. McLellan (University of Texas at Austin) graciously provided the mAb CR3022 and the spike protein expression vectors, and we thank the members of the Center for Systems and Synthetic Biology at the University of Texas at Austin for technical assistance. We thank Tom Anderson and Terumo BCT for continuously and rapidly supplying blood collection devices and supplies.

This study was supported by the National Institutes of Health grants AI146771-01 and AI139369-01, and the Fondren Foundation, Houston Methodist Hospital and Research Institute (to JMM). This research has been funded in whole or part with federal funds under a contract from the National Institute of Allergy and Infectious Diseases, National Institutes of Health, Contract Number 75N93019C00050 (to JL and GCI). A portion of this work was funded through Cooperative Agreement W911NF-12-1-0390 by the Army Research Office (to JDG).

